# Plasma protein profile associated with a family history of early-onset coronary heart disease

**DOI:** 10.1101/2024.10.29.24316396

**Authors:** Agnes Wahrenberg, Lars Lind, Natan Åberg, Henrike Häbel, Marika Ström, Anders Mälarstig, Patrik K.E. Magnusson, Ralf-Kuja Halkola, Göran Bergström, Gunnar Engström, Emil Hagström, Tomas Jernberg, Stefan Söderberg, Carl Johan Östgren, Per Svensson

**Affiliations:** Department of Clinical Science and Education, Södersjukhuset. Karolinska Institutet. Stockholm, Sweden; Department of Medical Sciences, Clinical Epidemiology, Uppsala University. Uppsala, Sweden; Department of Learning, Informatics, Management and Ethics, Karolinska Instituet. Stockholm, Sweden; Department of Medical Epidemiology and Biostatistics, Karolinska Institutet. Stockholm, Sweden; Department of Molecular and Clinical Medicine, Institute of Medicine, Sahlgrenska Academy, University of Gothenburg. Gothenburg, Sweden; Department of Clinical Physiology, Sahlgrenska University Hospital. Gothenburg, Sweden; Department of Clinical Sciences Malmö, Lund University. Malmö, Sweden; Department of Medical Sciences, Cardiology, Uppsala University. Uppsala, Sweden; Uppsala Clinical Research Center, Uppsala University. Uppsala, Sweden; Department of Clinical Sciences, Danderyd University Hospital, Karolinska Institutet. Stockholm, Sweden; Department of Public Health and Clinical Medicine, Medicine, Umeå University. Umeå, Sweden; Centre of Medical Image Science and Visualization, Linköping University. Linköping, Sweden; Department of Health, Medicine and Caring Sciences, Linköping University. Linköping Sweden.

## Abstract

**Background:** Proteins linked to heritable coronary heart disease (CHD) could uncover new pathophysiological mechanisms of atherosclerosis. We report on the protein profile associated with a family history of early-onset CHD and whether the relation between proteins and coronary atherosclerotic burden differs according to family history status, as well as inferences from mendelian randomization.

**Methods:** Data on coronary atherosclerotic burden from computed tomography angiography and Olink proteomics were retrieved for 4,521 subjects, free of known CHD, from the Swedish CArdioPulmonary bioImage Study (SCAPIS). Records of myocardial infarction and coronary revascularization therapies in any parent of subjects were retrieved from national registers. Linear associations between family history and proteins were adjusted for age, sex and study site. Statistical interactions between proteins and family history for the association between proteins and the coronary atherosclerotic burden were also studied. Mendelian randomization for causal associations between proteins and CHD was performed with GWAS summary data from UKB-PPP, CARDIoGRAMplusC4D and FinnGen.

**Results:** Of 4,251 subjects, family history of early-onset CHD was present in 9.5%. 38 proteins, with biological features of inflammation, lipid metabolism and vascular function, were associated with family history using a false discovery rate of 0.05. The strongest associations were observed for follistatin and cathepsin D, neither of which were attenuated by adjusting for cardiovascular risk factors.18 proteins were statistical interactors with family history in the association between each protein and the coronary atherosclerotic burden, most notably the LDL-receptor, transferrin receptor protein 1 and platelet endothelial cell adhesion molecule 1 (PECAM1). In two-sample mendelian randomization, a novel association was found for follistatin and myocardial infarction, and previous associations for PCSK9 and PECAM1 were repeated.

**Conclusions:** These findings highlight new potential mechanisms for heritable and general atherosclerosis.

**Clinical perspectives:** *What’s new?:*

- Proteins involved in inflammation and tissue remodeling, such as follistatin and cathepsin D, are strongly and independently associated with family history of early-onset CHD, indicating novel pathophysiological mechanisms of these proteins in familial disease.
- In subjects with a family history of early-onset CHD, the LDL-receptor, transferrin receptor protein 1 and PECAM1 were more strongly associated with coronary atherosclerotic burden, as compared to in subjects without family history.
- A novel, potentially causal association between follistatin and myocardial infarction was reported from Mendelian randomization using summary data from several GWAS projects.

*What are the clinical implications?:*

- This work brings evidence of the plasma protein profile in familial coronary heart disease, guiding further research of pathophysiological mechanisms in familial and general atherosclerotic disease.
- New target treatments for familial and general atherosclerotic disease may evolve from identified plasma proteins of importance in familial disease.

## Background

Having a family history of coronary heart disease (CHD) is a well-established risk factor for CHD, believed to represent the composite effect of familial genetic and environmental risk factors^1–3^. While a proportion of CHD heritability can be explained by risk factors such as dyslipidemias or CHD polygenic risk scores, a significant proportion is as of yet unexplained and may uncover new pathophysiological mechanisms in CHD. Recent advances in large-scale analysis of proteomics have led to new insights into mechanisms of atherosclerosis^4^. We sought to identify circulating proteins associated with a family history of early-onset CHD and to further study the relationship between identified proteins and coronary atherosclerotic burden in subjects with and without a family history of CHD. In addition, we aimed to identify causal roles of associated proteins in coronary atherosclerosis by using two-sample Mendelian randomization (MR). Lastly, we performed pathway enrichment analysis to explore biological themes across identified proteins.

## Methods

### Population

This was a study of the circulating plasma proteome in a middle-aged cohort from the Swedish CArdioPulmonary BioImage Study (SCAPIS)^5^. Individuals aged between 50 and 64 years were invited to participate in the prospective SCAPIS cohort, based on a random sample from the Swedish census register between 2013 and 2018, at six participating University sites in Sweden (Gothenburg, Linköping, Malmö/Lund, Stockholm, Umeå and Uppsala). The only exclusion criterion for SCAPIS was inability to understand spoken and written Swedish for the informed consent and study questionnaire. In total, 30,154 individuals were ultimately included in SCAPIS and underwent a comprehensive clinical examination, including physical anthropometry, health questionnaire, fasting blood sampling and coronary computed tomography angiography (CCTA). We included subjects from the SCAPIS cohort that gave explicit consent for linkage of their data to other registers, and that had fulfilled the CCTA protocol as well as the Olink biomarker analysis. We also excluded subjects reporting previous CHD if the form of myocardial infarction, percutaneous coronary intervention or coronary artery bypass grafting in the questionnaire. The implementation of the original SCAPIS study as well as subsequent analyses in this manuscript were granted by the Regional Ethical Board at Umeå University (2010-228-21M, 2017-183-31) and the Swedish Ethical Review Authority (2021-02951, 2022-04143-02).

### Clinical covariates

At the SCAPIS core clinical examination, body weight and height were measured with light clothing. Systolic blood pressure (SBP) was assessed with an automatic device, and the mean value of two measurements was reported in the study protocol. Smoking status was assessed in both an electronic questionnaire and with an oral question prior to lab sampling, classified as “Current”, “Former” and “Never” smoker. Previous medical history as well as usage of lipid-lowering and antihypertensive medications were also self-reported in the electronic questionnaire. A venous blood sample was drawn after an overnight fast for immediate analysis as well as for biobank storage for all SCAPIS participants. Analyses of total and low-density lipoprotein (LDL) cholesterol were made at the local lab of each participating site. For a subgroup of 5 075 participants, samples were sent for commercial proteomic biomarker analysis by Olink Proteomics (Uppsala, Sweden), with two pre-defined target panels comprising 184 biomarkers. The full scope of clinical examinations in SCAPIS has been described previously^5^.

### Coronary imaging

In study participants without contraindication to iohexol-contrast administration, an oral or intravenous beta-blocker as well as sublingual nitroglycerin was administered to study subjects with a heart rate above 60 beats/minute or a systolic blood pressure above 110 mmHg before scanning. Coronary imaging was performed using a dual-source computed tomography scanner (Somatom Definition Flash, Siemens Healthcare, Germany) with iohexol contrast medium at 325 mg I/kg body weight. Five different CCTA scanning protocols were used, chosen according to baseline coronary artery calcium score, heart rate and subject body weight. CCTA image sets were visually examined for coronary atherosclerosis by trained radiologists or cardiologists at the six participating sites and reported per segment using a model of l8 anatomical coronary segments^6^. The degree of coronary atherosclerosis was reported according to the degree of luminal stenosis as well as according to the presence of plaques, or non-assessable due to either the degree of plaque calcification or technical artefacts, for each coronary segment. A segment involvement score (SIS) was calculated as the sum of diseased coronary segments, regardless of the degree of luminal stenosis^7^.

### Analysis of circulating proteins

Analyses were performed by Olink Proteomics (Uppsala, Sweden), using two pre-defined Target panels (CVD II and CVD III), comprising 184 protein biomarkers selected by the vendor. Olink provides a proximity extension assay (PEA), where pairs of PEA probes with specific affinity for each target protein are equipped with an oligonucleotide sequence. When two probes bind to the target protein, the oligonucleotides hybridize and form a DNA template which is then amplified by a DNA polymerase and quantified using quantitative real-time polymerase chain reaction (qPCR). The qPCR readout for each target protein is proportional to the initial concentration in the sample. Biomarker qPCR readouts are provided as Normalised Protein eXpression (NPX) data on the Log2 scale, an arbitrary unit derived from qPCR cyclic threshold values that are compared to sample means from quality control markers in each sample plate. As the NPX value is normalized for each protein signal, high NPX values correspond to higher concentrations of the target protein, however, absolute NPX values cannot be compared across samples of different proteins.

### Register linkages and exposure definition

By means of unique personal identification numbers, issued to residents by the Swedish Tax Agency^8^, data on study participants were linked to national registers of kinship and diseases. First-degree relatives of study participants were identified in the Swedish Multi-Generation Register (MGR), containing information on relatives of a majority of individuals born in 1932 and later, resident in Sweden from January 1^st^ 1962 and onwards^9^. Data on relatives were linked to the National Patient Register and the Cause of Death Register, managed by the Swedish National Board of Health and Welfare, in order to identify registered manifestations of CHD in relatives. These registers include data on hospital admissions with registered main diagnoses according to the International Classification of Diseases (ICD) as well as registered causes of death according to ICD. Subjects missing information from any parent in the MGR were excluded from further analyses. Family history of early-onset CHD was defined as having at least one parent with a register-verified hospitalization or death due to a composite of either myocardial infarction or angina pectoris with any coronary revascularization procedure, occurring before the age of 55 years in male and 65 years in female relatives, as described previously^10^ and according to the European Society of Cardiology’s guidelines^11^. The corresponding ICD-10 and historical codes for CHD manifestations in relatives are provided in Supplementary Table S1.

### Statistical analysis in biomarker identification

Data management and statistical analyses were performed in STATA 16.1 (StataCorp, College Station, TX) and R (R version 4.3.1, The R Foundation). Baseline characteristics were tested for equality between individuals with and without family history of early-onset CHD with Pearsons’s chi-squared test for categorical variables, independent t-tests for normally distributed continuous variables and Mann-Whitney U test for non-normally distributed continuous variables. Associations between a family history of early-onset CHD and plasma levels of each protein were tested using linear regression with covariate adjustment for age, sex and study site. To account for multiple testing, the Benjamini-Hochberg method of a false discovery rate (FDR) was used. The Benjamini-Hochberg critical values were calculated using an FDR of 0.05, and a family history of early-onset CHD was considered to have a significant effect on a protein only if the regression coefficient’s p-value was lower than the respective critical value. It is henceforth noted as “at FDR<0.05” if this condition was met. Regression and correlation coefficients for associations significant at FDR<0.05 were illustrated in forest plots, volcano plots and heat maps. The model was then subsequently adjusted for other cardiovascular risk factors, including linearly for total cholesterol, LDL-cholesterol, systolic blood pressure and BMI, as well as for the use of lipid-lowering or antihypertensive medication, smoking status, and presence of diabetes mellitus. Similarly, the linear association between each biomarker and SIS were assessed with linear regression, adjusted for age, sex and study site in a basic model and in an adjusted model with other cardiovascular risk factors. Additionally, a third model included statistical interaction terms between family history of early-onset CHD and each biomarker. The association between biomarkers and SIS was also examined in subgroups of family history status. Lastly, the association between a family history of early-onset CHD and SIS was examined with linear regression.

### Two-sample Mendelian randomization for CHD

A two-sample Mendelian randomization (MR) approach was used to explore possible causal relationships between biomarkers and CHD, to further examine the pathophysiology of heritable CHD. For the proteins that were significantly associated with family history of early-onset CHD or that were among the top 20 proteins associated with SIS in subjects with positive family history, or those that were significant statistical interactors in the relationship between family history and SIS, we systematically searched for genetic instrumental variables (IV) for MR. Protein quantitative trait loci (pQTL) for each of the selected proteins, with an F-statistic of at least 50, were identified from the UK Biobank Pharma Proteomics Project (UKB-PPP), in which GWAS for an abundance of circulating proteins has been performed in 54,219 individuals^12^. We used only the sentinel cis-pQTL with the lowest p-value (p < 1.7*10^−11^) from the complete UKB-PPP cohort for each protein as IVs to reduce the risk of horizontal pleiotropy. For proteins of which the sentinel cis-pQTL was a multiallelic variant in the complete cohort, the meta-analysis statistic from the discovery and replication cohort was used if it was a biallelic variant. If the cis-pQTL was also multiallelic in the discovery and replication cohorts, a biallelic SNP with the lowest p-value within a 1 Mb window of the coding gene from the combined cohort was used. Outcome data on coronary heart disease and myocardial infarction from the CARDIoGRAMplusC4D 1000 Genomes project have been contributed by CARDIoGRAMplusC4D investigators and have been downloaded from www.CARDIOGRAMPLUSC4D.ORG, including 60,801 cases and 123,504 controls^13^. Further, outcome data on myocardial infarction were also retrieved from the Finngen R10 populations study release, including 26,060 cases and 360,108 controls^14^. The Wald ratio was used to calculate a causal estimate of the association between each protein and myocardial infarction. Causal estimates of association between genetic determinants of proteins and myocardial infarction from CARDIoGRAMplusC4D and Finngen R10 outcome data were subsequently combined in meta-analysis using a fixed-effects model. The limit of significance was Bonferroni adjusted to *p= 0,001389*. The pQTLs included as IVs in the analysis are provided in Supplementary Table S2.

### Analysis of biological pathways

To investigate biological pathways and processes relevant to heritable coronary atherosclerosis we performed exploratory analyses in g:Profiler^15^, studying whether there were signs of overrepresentation of biological terms in the genes coding for the 38 proteins associated with a family history of early-onset CHD at FDR<0.05. For this gene set, we performed an unordered enrichment analysis of biological terms in Kyoto Encyclopedia of Genes and Genomes (KEGG)^16^, Reactome (REAC)^17^; Wiki Pathways (WP)^18^ and Gene Ontology (GO)^19,20^. Thereafter, we compared overrepresented pathways as mirrored by the different proteomic profiles of coronary atherosclerosis in those with and without early-onset family history of early-onset CHD, using the 20 proteins most strongly associated with coronary atherosclerotic burden in the two groups. For this analysis we performed a similar unordered enrichment analysis as above using a multiquery to compare the two gene sets.

## Results

### Biomarker identification

In total, 4,251 subjects free of self-reported CHD were included from SCAPIS. A family history of early-onset CHD was present in 405 (9.5%) subjects. The exclusion process is visualized in Figure 1. Baseline characteristics according to family history status are presented in Table 1. Subjects with a family history of early-onset CHD were younger and more commonly female. The levels of LDL-cholesterol were largely the same across family history groups, however, family history positive subjects were more often on a lipid-lowering therapy which could explain the lack of differences in LDL-cholesterol levels. Descriptives of subjects excluded due to known CHD are presented in Supplementary Table S3.

**Figure 1.**
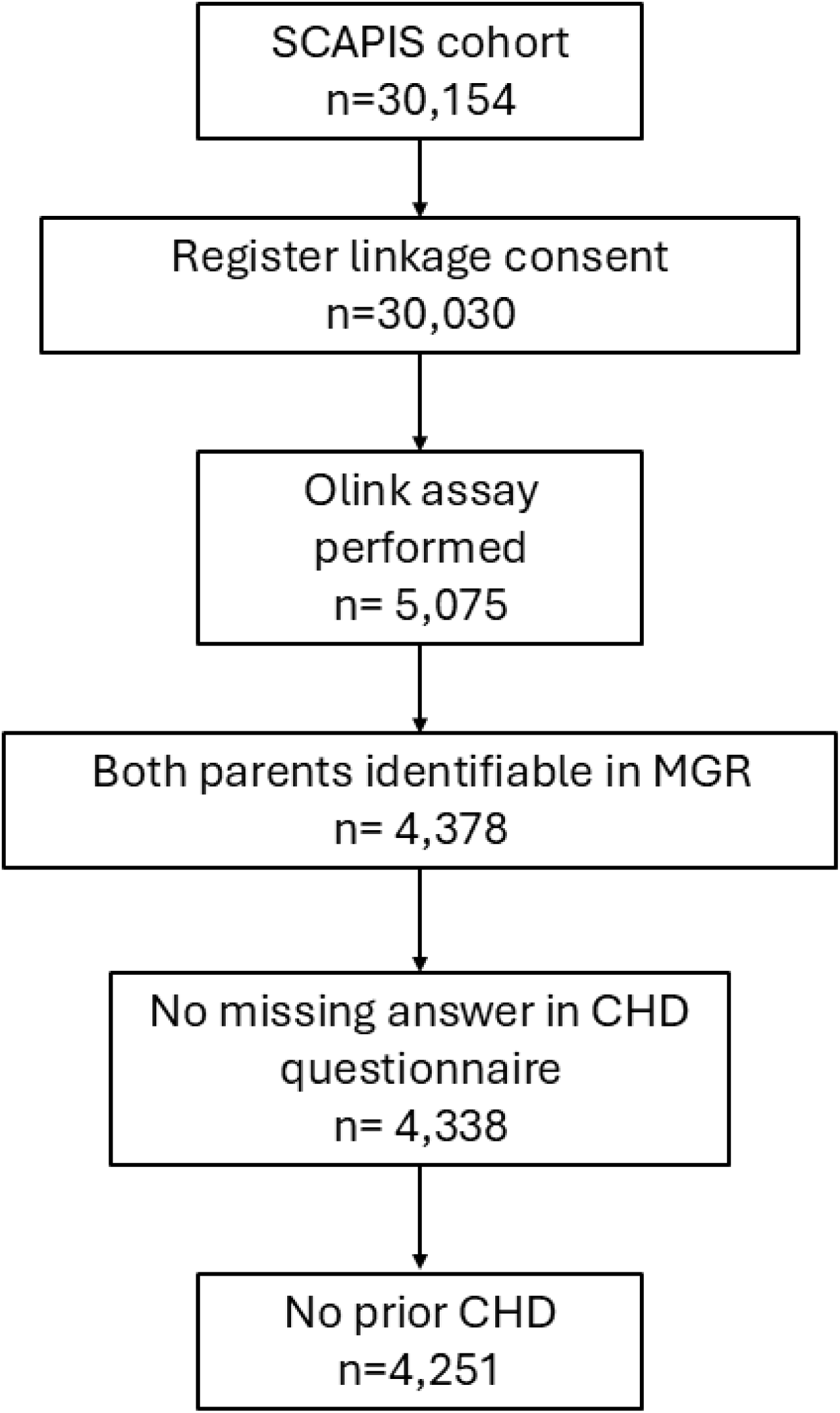
Number of patients throughout the exclusion process. Abbreviations: CHD – coronary heart disease. MGR – the Swedish Multi-Generation Register. SCAPIS – Swedish CArdioPulmonary BioImage Study.

**Table 1.** Subject characteristics according to family history status.

|  | **Family history of**<br>**early-onset CHD**<br>**(n=405)** | **No family history of**<br>**early-onset CHD**<br>**(n=3,846)** | ***p-value*** |
| --- | --- | --- | --- |
| **Female** | 233 (57.5%) | 1,945 (50.6%) | 0.008 |
| **Age** | 57.1 (4.3) | 57.6 (4.3) | 0.050 |
| **Site** |  |  | 0.13 |
| **Göteborg** | 62 (15.3%) | 626 (16.3%) |  |
| **Linköping** | 88 (21.7%) | 678 (17.6%) |  |
| **Malmö** | 61 (15.1%) | 574 (14.9%) |  |
| **Stockholm** | 56 (13.8%) | 607 (15.8%) |  |
| **Umeå** | 82 (20.2%) | 689 (17.9%) |  |
| **Uppsala** | 56 (13.8%) | 672 (17.5%) |  |
| **Smoking status** |  |  | 0.82 |
| **Former smoker** | 143 (35.3%) | 1,413 (36.7%) |  |
| **Never smoked** | 207 (51.1%) | 1,968 (51.2%) |  |
| **Current smoker** | 49 (12.1%) | 410 (10.7%) |  |
| **Missing** | 6 (1.5%) | 55 (1.4%) |  |
| **History of diabetes** | 27 (6.7%) | 127 (3.3%) | <0.001 |
| **BMI (kg/m<sup>2</sup>)** | 27.17 (4.57) | 26.59 (4.16) | 0.008 |
| **HDL cholesterol (mmol/l)** | 1.6 (0.49) | 1.7 (0.49) | 0.38 |
| **LDL cholesterol (mmol/l)** | 3.4 (0.98) | 3.4 (0.94) | 1.00 |
| **Total cholesterol (mmol/l)** | 5.6 (1.08) | 5.6 (1.05) | 0.75 |
| **Triglycerides (mmol/l)** | 1.2 (0.43) | 1.1 (0.39) | 0.001 |
| **Lipid lowering medication** | 53 (13.1%) | 266 (6.9%) | <0.001 |
| **Antihypertensive medication** | 93 (23.0%) | 728 (18.9%) | 0.050 |
| **Systolic blood pressure (mmHg)** | 129 (16.7) | 127 (16.7) | 0.11 |
| **Segment involvement score**<br>**(SIS)** | 1.56 (2.23) | 1.05 (1.78) | <0.001 |
| **Moderate to severe coronary**<br>**atherosclerosis (SIS<sub>≥</sub>4)** | 83 (20.5) | 379 (9.8) | < 0.001 |

Values are N (%) or mean (SD). Abbreviations: BMI, body mass index. HDL, high-density lipoprotein. LDL, low-density lipoprotein.

In a basic model adjusted for age, sex and study site, a total of 38 biomarkers were significantly associated with family history of early-onset CHD using at FDR<0.05, presented in Figure 2 and Table 2. All protein associations with family history of early-onset CHD are listed in Supplementary Table S4. After adjusting for cardiovascular risk factors, 27 biomarkers remained significantly associated with family history of early-onset CHD at p<0.05. The correlation of individual proteins and cardiovascular risk factors are illustrated in a heatmap in Figure 2. Paraoxonase (PON3), stem cell factor (SCF) and lipoprotein lipase (LPL) were negatively associated with family history of early-onset CHD, whereas some of the biomarkers with the strongest positive correlation with family history were cathepsin D (CTSD), renin (REN), Interleukin-1 receptor antagonist protein (IL-1RA) and follistatin.

**Figure 2.**
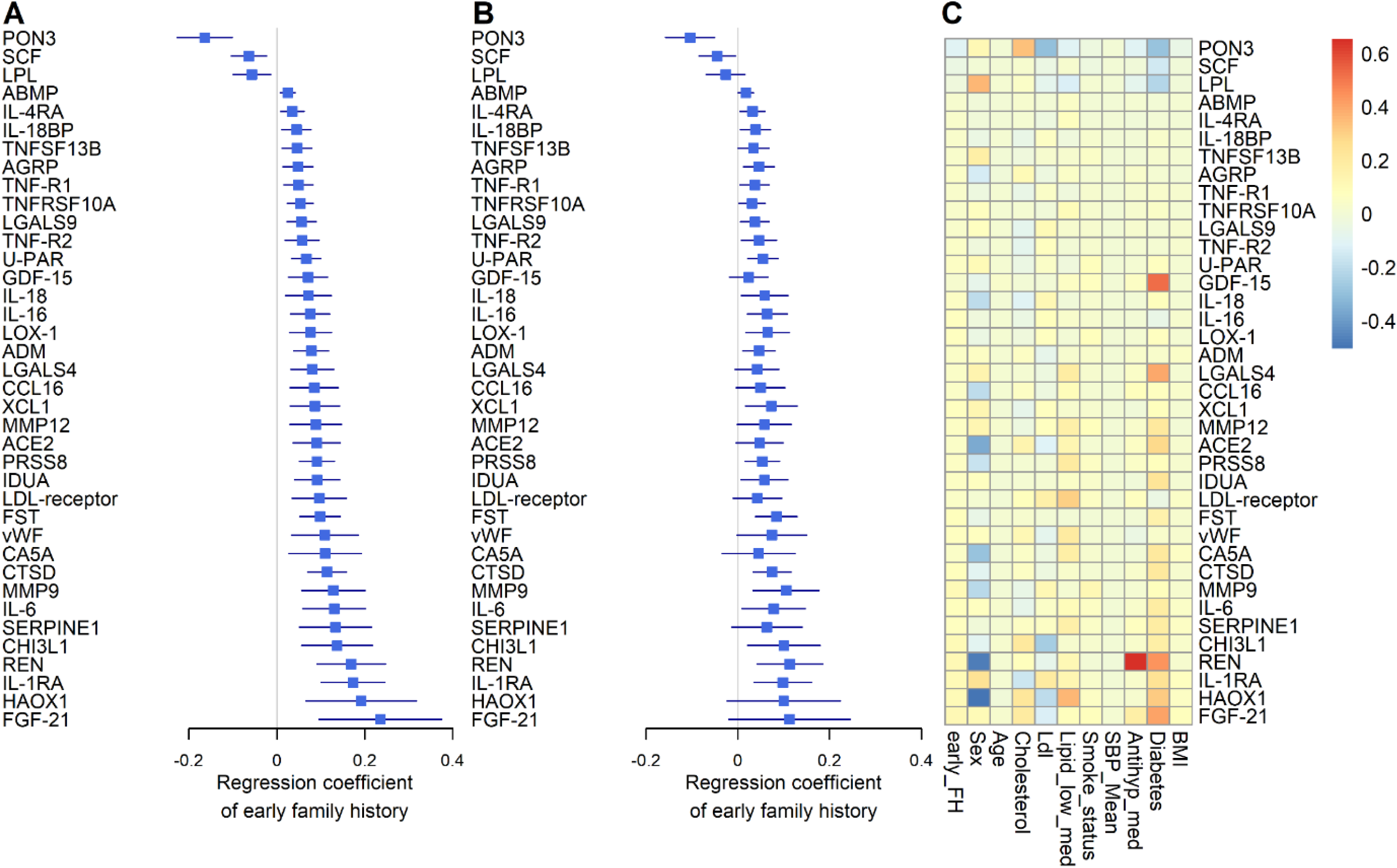
Proteins significantly associated with family history of early-onset CHD. (A) Regression coefficients and 95% confidence intervals (CI) of the proteins associated with family history, adjusted for age, sex and study site, significant using a false discovery rate of <0.05. (B) Regression coefficients and 95% CIs of proteins significantly associated with family history, additionally adjusted for body mass index (BMI), diabetes, antihypertensive treatment, mean systolic blood pressure (SBP), smoking status, lipid lowering treatment, LDL-cholesterol and total cholesterol. (C) Heat map of the correlation of proteins with each cardiovascular risk factor.

**Table 2.**
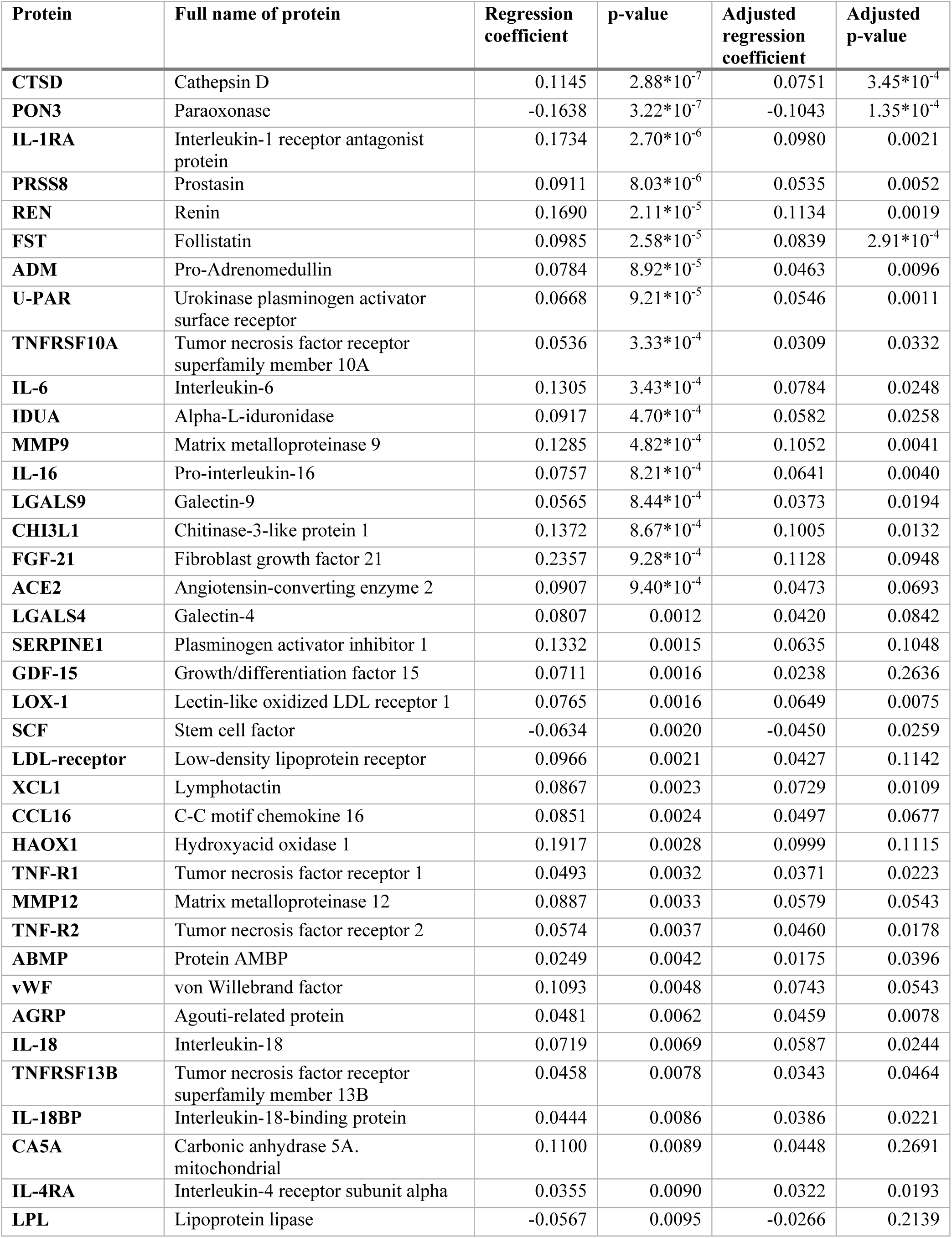
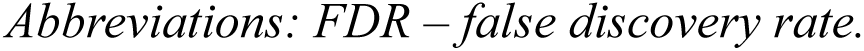
Proteins significantly associated with family history of early-onset coronary heart disease at FDR<0.05.

| Protein | Full name of protein | Regression coefficient | p-value | Adjusted regression coefficient | Adjusted p-value |
| --- | --- | --- | --- | --- | --- |
| **CTSD** | Cathepsin D | 0.1145 | $2.88 \times 10^{-7}$ | 0.0751 | $3.45 \times 10^{-4}$ |
| **PON3** | Paraoxonase | -0.1638 | $3.22 \times 10^{-7}$ | -0.1043 | $1.35 \times 10^{-4}$ |
| **IL-1RA** | Interleukin-1 receptor antagonist protein | 0.1734 | $2.70 \times 10^{-6}$ | 0.0980 | 0.0021 |
| **PRSS8** | Prostasin | 0.0911 | $8.03 \times 10^{-6}$ | 0.0535 | 0.0052 |
| **REN** | Renin | 0.1690 | $2.11 \times 10^{-5}$ | 0.1134 | 0.0019 |
| **FST** | Follistatin | 0.0985 | $2.58 \times 10^{-5}$ | 0.0839 | $2.91 \times 10^{-4}$ |
| **ADM** | Pro-Adrenomedullin | 0.0784 | $8.92 \times 10^{-5}$ | 0.0463 | 0.0096 |
| **U-PAR** | Urokinase plasminogen activator surface receptor | 0.0668 | $9.21 \times 10^{-5}$ | 0.0546 | 0.0011 |
| **TNFRSF10A** | Tumor necrosis factor receptor superfamily member 10A | 0.0536 | $3.33 \times 10^{-4}$ | 0.0309 | 0.0332 |
| **IL-6** | Interleukin-6 | 0.1305 | $3.43 \times 10^{-4}$ | 0.0784 | 0.0248 |
| **IDUA** | Alpha-L-iduronidase | 0.0917 | $4.70 \times 10^{-4}$ | 0.0582 | 0.0258 |
| **MMP9** | Matrix metalloproteinase 9 | 0.1285 | $4.82 \times 10^{-4}$ | 0.1052 | 0.0041 |
| **IL-16** | Pro-interleukin-16 | 0.0757 | $8.21 \times 10^{-4}$ | 0.0641 | 0.0040 |
| **LGALS9** | Galectin-9 | 0.0565 | $8.44 \times 10^{-4}$ | 0.0373 | 0.0194 |
| **CHI3L1** | Chitinase-3-like protein 1 | 0.1372 | $8.67 \times 10^{-4}$ | 0.1005 | 0.0132 |
| **FGF-21** | Fibroblast growth factor 21 | 0.2357 | $9.28 \times 10^{-4}$ | 0.1128 | 0.0948 |
| **ACE2** | Angiotensin-converting enzyme 2 | 0.0907 | $9.40 \times 10^{-4}$ | 0.0473 | 0.0693 |
| **LGALS4** | Galectin-4 | 0.0807 | 0.0012 | 0.0420 | 0.0842 |
| **SERPINE1** | Plasminogen activator inhibitor 1 | 0.1332 | 0.0015 | 0.0635 | 0.1048 |
| **GDF-15** | Growth/differentiation factor 15 | 0.0711 | 0.0016 | 0.0238 | 0.2636 |
| **LOX-1** | Lectin-like oxidized LDL receptor 1 | 0.0765 | 0.0016 | 0.0649 | 0.0075 |
| **SCF** | Stem cell factor | -0.0634 | 0.0020 | -0.0450 | 0.0259 |
| **LDL-receptor** | Low-density lipoprotein receptor | 0.0966 | 0.0021 | 0.0427 | 0.1142 |
| **XCL1** | Lymphotactin | 0.0867 | 0.0023 | 0.0729 | 0.0109 |
| **CCL16** | C-C motif chemokine 16 | 0.0851 | 0.0024 | 0.0497 | 0.0677 |
| **HAOX1** | Hydroxyacid oxidase 1 | 0.1917 | 0.0028 | 0.0999 | 0.1115 |
| **TNF-R1** | Tumor necrosis factor receptor 1 | 0.0493 | 0.0032 | 0.0371 | 0.0223 |
| **MMP12** | Matrix metalloproteinase 12 | 0.0887 | 0.0033 | 0.0579 | 0.0543 |
| **TNF-R2** | Tumor necrosis factor receptor 2 | 0.0574 | 0.0037 | 0.0460 | 0.0178 |
| **ABMP** | Protein AMBP | 0.0249 | 0.0042 | 0.0175 | 0.0396 |
| **vWF** | von Willebrand factor | 0.1093 | 0.0048 | 0.0743 | 0.0543 |
| **AGRP** | Agouti-related protein | 0.0481 | 0.0062 | 0.0459 | 0.0078 |
| **IL-18** | Interleukin-18 | 0.0719 | 0.0069 | 0.0587 | 0.0244 |
| **TNFRSF13B** | Tumor necrosis factor receptor superfamily member 13B | 0.0458 | 0.0078 | 0.0343 | 0.0464 |
| **IL-18BP** | Interleukin-18-binding protein | 0.0444 | 0.0086 | 0.0386 | 0.0221 |
| **CA5A** | Carbonic anhydrase 5A. mitochondrial | 0.1100 | 0.0089 | 0.0448 | 0.2691 |
| **IL-4RA** | Interleukin-4 receptor subunit alpha | 0.0355 | 0.0090 | 0.0322 | 0.0193 |
| **LPL** | Lipoprotein lipase | -0.0567 | 0.0095 | -0.0266 | 0.2139 |

*Abbreviations: FDR – false discovery rate.*

Mean SIS was 1.56 (±2.23 SD) in subjects with a family history of early-onset CHD, and 1.04 (±1.78 SD) in subjects without family history. The distribution of SIS scores is shown in Supplementary Table S5. Family history of early-onset CHD was significantly associated with SIS, with an age, sex and site-adjusted SIS of 0.64 units higher in subjects with family history as compared to those without family history (95% CI 0.46 – 0.81, p = 1.4*10^−12^).

Overall, 79 biomarkers were associated with SIS in a basic model at FDR<0.05 out of which 28 were also associated with a family history of early-onset CHD. The associations between each protein and SIS were also modelled in subgroups according to family history status, presented in Figure 3 and Supplementary Table S6. In subjects with family history of early-onset CHD, a strong negative association with SIS was observed for PON3, whereas strong positive associations were observed for LDL-receptor, matrix metalloproteinase 12 (MMP12) and tissue-type plasminogen activator (t-PA). There was also a positive relationship between transferrin receptor protein 1 (TR) and SIS in subjects with family history, whereas the association was non-significant with a negative coefficient for subjects without family history. Notably, among the proteins associated with family history of early-onset CHD, some were however not significantly associated with SIS in subjects with positive family history, such as SCF, LPL, REN, and growth/differentiation factor 15 (GDF-15). The 20 proteins with the strongest association with SIS at FDR<0.2 in each subgroup are shown in Supplementary Figure S1.

**Figure 3.**
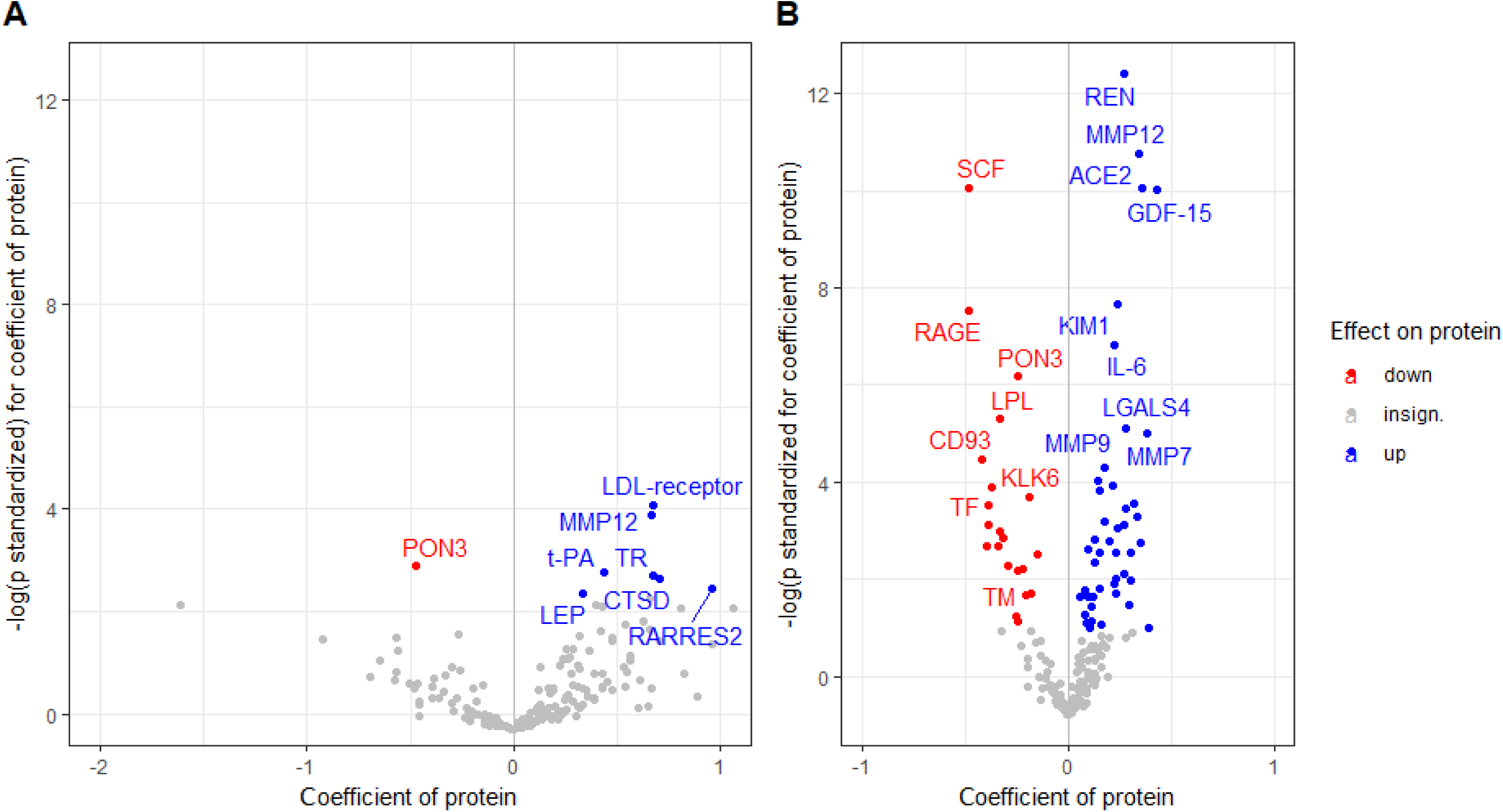
Volcano plot of regression coefficients and probability values for the associations between proteins and the segment involvement score using a false discovery rate of <0.05. Panel A depicts proteins significant among subjects with a family history of early CHD, whereas panel B depicts proteins significant among subjects without such family history.

In order to test whether the associations between circulating proteins and coronary atherosclerotic burden differed between subjects according to family history status, interaction tests between family history and each protein in the association between proteins and SIS were performed. Significant statistical interactions were observed for 18 proteins at p<0.05, shown in Figure 4, Supplementary Figure S2 and Supplementary Table S7. In subjects with family history, Interleukin-17D (IL_17D), LPL and PON3 had a steeper inverse association with SIS, whereas circulating LDL-receptor, TR, tissue-type plasminogen activator (T-PA), CTSD, cathepsin Z (CTSZ) and platelet endothelial cell adhesion molecule (PECAM1) showed a steeper positive association with SIS.

**Figure 4.**
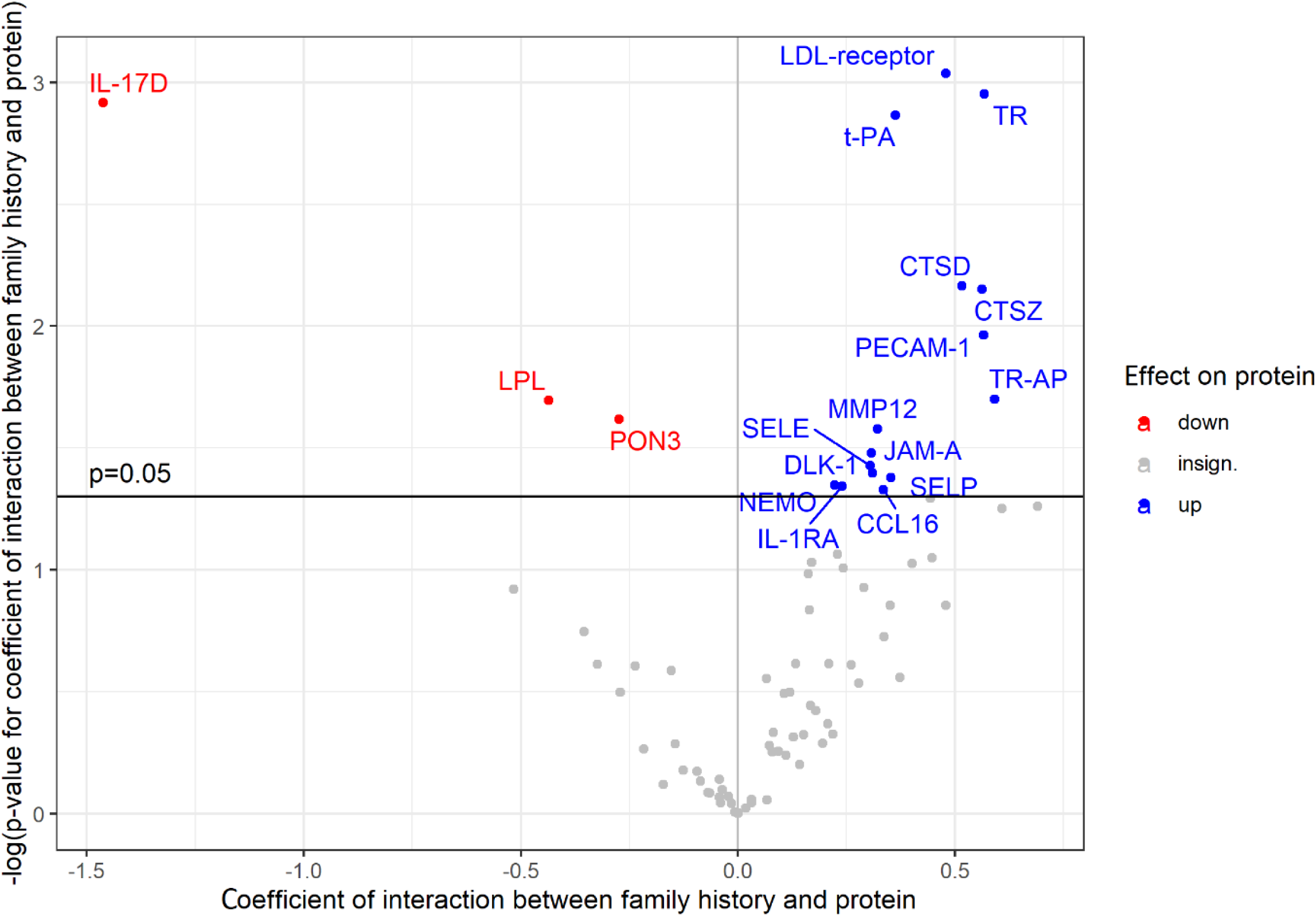
Volcano plot of interaction coefficients and probability values for the interaction between family history of early coronary heart disease and each protein in the associations between proteins and the segment involvement score.

### Two-sample MR for CHD

Proteins that were associated with family history of early-onset CHD, or among the top 20 proteins most strongly associated with SIS in subjects with family history, or that were significant interactors with family history in the relationship with SIS were selected for two-sample MR. In total, 36 proteins that had strong genetic instruments available in data from the UKB-PPP^12^ were used in the analysis. At a Bonferroni adjusted p<0,001389, three proteins showed potential causal associations with myocardial infarction in the meta-analysed MR. MR odds ratios for myocardial infarction are shown in Figure 5 and Supplementary Table S8. PECAM1, proprotein convertase subtilisin/kexin type 9 (PCSK9) and follistatin showed evidence of causality, of which PECAM1 conferred the greatest combined OR for CHD of 2.52 (95% CI 1.81 – 4.00, *p=1.57×10*^−6^). We also noted a trend towards significance for additional proteins, such as LPL and MMP12, both with negative directions of effect in relation to myocardial infarction.

**Figure 5.**
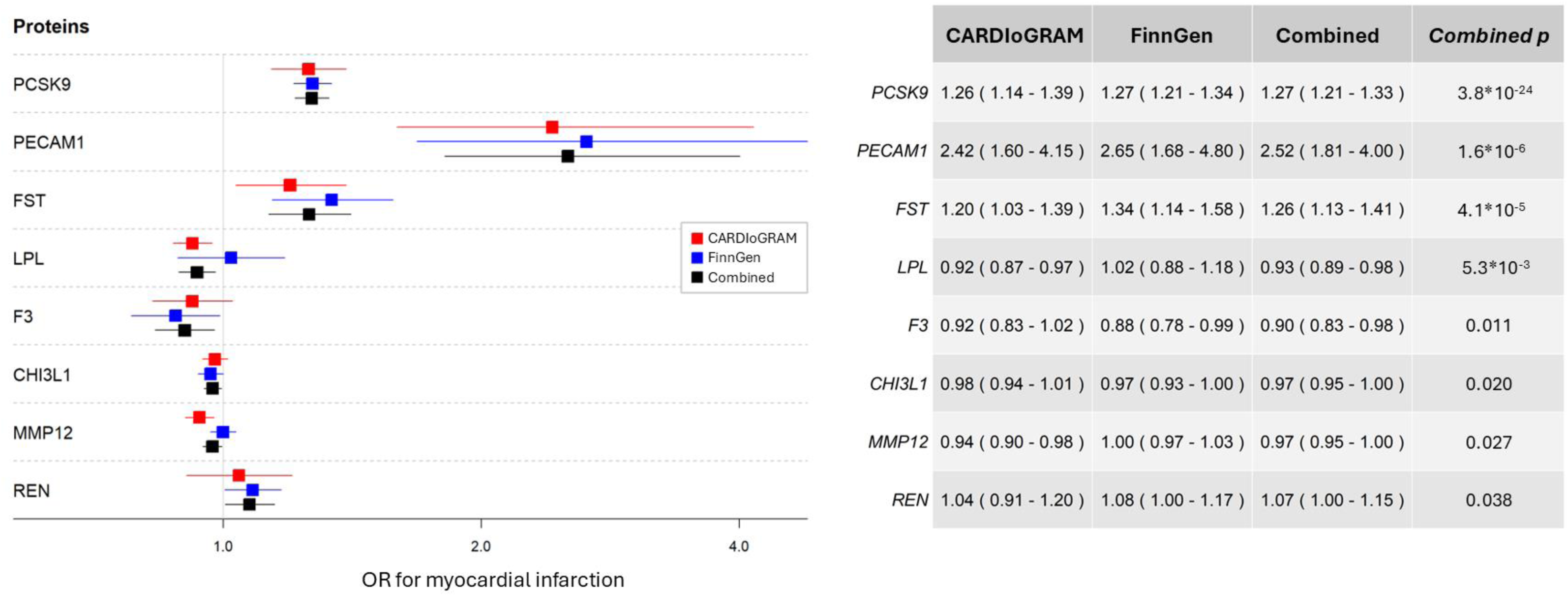
Mendelian randomization odds ratios for selected proteins and myocardial infarction. Odds ratios (OR) and 95% CIs for every genetically predicted ±SD of the normalized protein level on myocardial infarction, for the 8 strongest associations of tested proteins. Results shown for outcome data from CARDIoGRAM, FinnGen and the meta-analysis, respectively.

### Pathways associated with family history of early-onset CHD and heritable coronary atherosclerosis

The biological terms that were identified in an exploratory pathway enrichment analysis are presented in Supplementary Figures S3 and S4. The following biological pathways were the ones most highly enriched in subjects with family history of early-onset CHD: *inflammatory response* (GO:0006954) and *response to organic substance* (GO:0010033). Augmented molecular functions were *signaling receptor binding* (GO:0005102) and *death receptor activity* (GO:0005035) and enriched cellular components were *extracellular region* (GO:0005615) and *extracellular matrix* (GO:0031012, as expected when measuring circulating plasma proteins. Different biological pathways were enriched when the proteomic profile associated with coronary atherosclerotic burden was compared between those with and without a family history of early-onset CHD. Using GO:BP, in heritable coronary atherosclerosis, *inflammatory response* (GO:0006354) was more enriched in those with as compared to those without family history. The molecular function *lipoprotein particle binding* (GO:0071813) was clearly augmented in subjects with family history as compared to those without. Among cellular components, the *PCSK9-LDLR complex* (GO:1990666) was enriched in subjects with family history as compared to those without family history.

## Discussion

In this paper, we identify circulating proteins associated with a family history of early-onset CHD and describe how these proteins are associated with coronary atherosclerotic burden in subjects of the general population, free of previously reported CHD. Further, we study how a family history of early-onset CHD may influence the way in which these proteins associate with coronary atherosclerotic burden. By using two-sample MR, we report potential causal effects of several proteins on the risk of myocardial infarction, a detrimental manifestation of CHD. We also explore the biological pathways that may be involved in hereditary CHD. In summary, out of 184 tested proteins 38 were significantly associated with a family history of early-onset CHD and 79 with the coronary atherosclerotic burden. Furthermore, a significant statistical interaction with family history was observed for 18 proteins in their association with the coronary atherosclerotic burden, with steeper positive associations between circulating LDL-receptor, TR, tissue-type plasminogen activator (T-PA) and coronary atherosclerosis in those with family history of early-onset CHD. We establish follistatin as a novel addition to proteins involved in familial coronary atherosclerosis, and in a two-sample MR analysis, for the first time we report a potential causal relationship between follistatin and myocardial infarction. Moreover, we confirm previous reports on causal effects of proteins involved in lipid metabolism and the risk of myocardial infarction, for PCSK9 and PECAM1. Our findings give new insights into the pathophysiological mechanisms of heritable CHD, as the identified proteins may contribute to new prediction tools or treatment targets.

Since decades CHD has been known as a familial disease^21^, and although metabolic risk factors may aggregate within families, independent familial effects on CHD risk have repeatedly been reported^22^. Serum lipid levels are highly heritable, and elevated LDL-cholesterol is established as a causal factor in CHD pathophysiology^23,24^. Elevated circulating PCSK9-levels escalate LDL-receptor degradation and thus increase circulating LDL-cholesterol. We confirm the results of previous MR-studies, showing a causal effect of PCSK9 on the risk of myocardial infarction^25^. Furthermore, levels of soluble LDL-receptor were strongly associated with atherosclerotic burden, as measured by SIS, in subjects with a family history of early-onset CHD, and was also an interactor with family history in the relationship between soluble LDL-receptor and SIS. While there was no strong genetic instrument for the soluble LDL-receptor in our MR analysis to estimate causal effects, our findings further underline the role of LDL-turnover in the development of heritable CHD. LPL was negatively associated with family history of early-onset CHD, and in subjects with family history there was a steeper negative relationship between LPL and coronary atherosclerotic burden as compared to subjects without family history. In MR, LPL showed a trend towards an inverse causal relationship with CHD, in line with previous studies indicating a higher risk of CHD in subjects with loss-of-function variants of LPL as well as a protective relationship between triglyceride-lowing LPL variants and CHD in MR analyses^26^.

PON3 inhibits LDL-oxidation and has been shown to slow atherosclerosis progression^27^. In this study, PON3 was strongly negatively associated with both family history of CHD and SIS, however, not meeting the corrected significance level in our MR analysis. Nevertheless, the negative associations between PON3 and family history as well as coronary atherosclerotic burden were strong and independent of traditional cardiovascular risk factors, accentuating the importance of LDL-oxidation in CHD development, as well as the importance of familial factors influencing such oxidation.

PECAM1 is involved in cell adhesion and the patency of the vascular endothelial barrier, as well as in signaling between endothelial cells and migrating cells involved in inflammation and hemostasis. Increased PECAM1-levels in response to shear stress of the vessel wall has been associated with the development of atherosclerotic plaques in lesion-prone regions of the aorta in mice^28^, in which an elevated expression of PECAM1 locally in the vessel wall of such aortic section. In this study, PECAM1 exhibited a steeper association curve with SIS in subjects with family history of CHD and showed a causal, positive relationship with myocardial infarction in MR. However, PECAM1 was not significantly associated with a family history of early-onset CHD, nor the coronary atherosclerotic burden as measured by SIS. This suggests that while a family history of CHD is not necessarily predictive of PECAM1-levels, there may be modulating effects on PECAM1 functions. Additionally, as PECAM1 was not associated with coronary atherosclerotic burden in subjects without previously known CHD in this study, there may instead be modulating effects of PECAM1 in late-stage atherosclerosis, possibly inflected by other familial factors. We replicate the findings of Lind et al.^29^, reporting an association between PECAM1 and myocardial infarction from two-sample Mendelian randomization, along with strong support of causal variants shared between PECAM1 and CHD in colocalization analyses.

Follistatin was positively associated with a family history of early-onset CHD, independent of traditional risk factors, and associated with coronary atherosclerotic burden in the overall analysis of all subjects. The results from two-sample MR further suggest that follistatin may play a causal role in CHD, and potentially contribute to the development of heritable coronary atherosclerosis. Follistatin is known to modulate the inflammatory response and muscle growth signaling, indicating a role in tissue repair and remodeling. Our findings suggest potential familial effects on follistatin expression or function, as well as a role of follistatin in the development of atherosclerosis. Circulating follistatin has previously been linked to mortality due to heart failure and stroke, partly mediated through diabetes mellitus^30^. Previous MR analyses have established a potential causal association between follistatin and diabetes mellitus^31^, however, to our knowledge no causal relationship between follistatin and CHD has previously been published until now.

Whereas not meeting the Bonferroni corrected *p* for causality in the MR analysis, several proteins involved in inflammation exhibited intriguing associations with family history. MMP-12, a regulator of vascular macrophage infiltration, has previously been positively associated with heart failure, ischemic stroke and CHD^32,31^, and was associated with both family history of early-onset CHD and SIS in this study. However, our MR findings were borderline indicative of an inverse causal relationship between MMP-12 and CHD. This has been observed previously in works of Lind et al^31^, where MMP-12 was positively associated with ischemic stroke and CHD, while exhibiting an inverse causal association in MR analysis. Similarly, MMP-12 has been positively associated with peripheral artery disease, while in the same material, a negative causal relationship between MMP-12 and peripheral artery disease was observed in MR^33^. Reverse MR by Li et al^34^ showed signs of reverse causality of CVD on MMP-12, indicating a possible feedback mechanism of CHD on MMP-12 expression. The clinical significance of this potential feedback mechanism is still unknown.

CTSD is a regulator of lysosomal degradation and elevated levels in plasma and myocardial cells has been associated with myocardial injury and coronary events^35,36^. It has been suggested that dysfunctional autophagy may lead to atherosclerotic plaque formation, and that advanced atherosclerosis may lead to overexpression of CTSD, as autosomal systems cannot manage the burden of oxidative stress. In this study CTSD, similarly as for MMP-12, was strongly connected to both family history of early-onset CHD and overall atherosclerotic burden. CTSD also demonstrated a steeper association curve with SIS in subjects with positive family history of early-onset CHD. However, a causal association between CTSD and CHD was not found in MR, which may emphasize the theory that CTSD is elevated as a response to extensive oxidative stress rather than causative of atherosclerosis.

C-C motif chemokine 16 (CCL16) is a chemotactic agent found in plasma and liver tissue involved in inflammatory response, however, little is known about its role in disease development. In this study, CCL16 was positively associated with family history of early-onset CHD, as well as with overall SIS, and was an interactor with family history in the association with SIS. However, no evidence of a causal association with CHD was found. Previously, CCL16 has been associated with incident myocardial infarction^37^ and microvascular dysfunction in non-obstructive angina in women^38^, however, the mechanism of CCL16 in coronary heart disease is still unclear.

The inverse association between IL-17D and coronary atherosclerotic burden in this study was significantly steeper in subjects with a family history of early-onset CHD. The interleukin-17 family has been proposed to be involved in coronary atherosclerosis, reportedly associated with instability of coronary plaques^39^. IL-17D has also been associated with all-cause mortality in patients with heart failure^40^, however, the pathophysiological importance of each member of the interleukin-17 family in atherosclerosis remains unsettled.

Soluble TR is upregulated in iron deficiency. We have shown that TR is associated with coronary atherosclerotic burden in subjects with family history of early-onset CHD, whereas it is not associated with SIS in subjects without such history. Thereto, TR was a potent interactor with family history in the association with SIS. It has been suggested that relative iron deficiency, even before hemoglobin levels are afflicted, may influence the inflammation, remodeling and survival of cardiomyocytes in myocardial infarction and iron deficiency as measured with the soluble TR has been associated with cardiovascular mortality and myocardial infarction in subjects with known CHD^41^. Additionally, TR has been associated with an increased risk of aortic valve replacement due to artic stenosis, only in subjects with established CHD^42^. Our findings highlight that iron metabolism may play a particular role in the development of heritable CHD.

### Strengths and limitations

The greatest strength of this study is in the unique way that data on proteins, cardiovascular risk factors and CCTA-verified coronary atherosclerotic burden in the general population are combined with national registers of kinship and diseases. This increases the robustness of disease history, particularly for early manifestations of familial disease. Furthermore, we included data from international GWAS datasets to infer causal effects of proteins related to heritable CHD on coronary atherosclerotic burden, triangulating evidence from different methods. The meta-analysis of GWAS datasets improves the power to infer such causal associations. There are, however, limitations pertinent to the inference of this study. Firstly, due to the unique features of the SCAPIS cohort that combines cardiac imaging and proteomic data in a population-based setting, and the registers that enable the identification of register-based familial disease, there is no similar cohort available for replication. Due to the limited number of subjects in the SCAPIS study undergoing testing for proteomic markers, the cohort was too small to purposefully divide into separate discovery and validation cohorts. Secondly, the strength of the genetic instruments used in MR is the determining factor of finding causal effects of proteins on a designated outcome, in this case myocardial infarction. Including several instruments per protein would further increase power to find proteins causally involved in atherosclerosis. However, we used only the strongest pQTL available as the IV for each protein, increasing robustness and strength of instruments, while also reducing the risk of horizontal pleiotropy pertinent to including several weaker variants in IVs. Thirdly, the inclusion of additional cis-pQTLS as well as trans-pQTLs could possibly identify other proteins causally associated with CHD, for which we did not find strong enough instruments with our approach, but could also increase the risk of horizontal pleiotropy and thus compromising the core assumption of exclusion restriction of MR.

## Conclusion

In this study we identify several plasma proteins strongly associated with family history of early-onset CHD with biological features of inflammation, lipid metabolism and vascular function. Further, family history of early-onset CHD strongly influenced how different proteins associated with the coronary atherosclerotic burden, suggesting a specific proteomic profile of heritable coronary atherosclerosis. Notably, through MR analysis we have established a possible causal relationship between PCSK9, PECAM1 and follistatin and myocardial infarction, which may have implications for further understanding of the pathophysiology of CHD.

## Data Availability

Due to the sensitive information of studied subjects and identified relatives and according to the decision of the Swedish Ethical Review Authority, data will not be made publicly available.

